# EXPLORING THE PHYSICAL WELL-BEING OF BREAST CANCER PATIENTS IN KUMASI METROPOLIS: A QUALITATIVE STUDY

**DOI:** 10.1101/2024.05.01.24306685

**Authors:** Victoria Sefah, Hayford Isaac Budu, Priscilla Felicia Tano, Emmanuel Kofi Lawer, Emile Kouakou Tano

**Affiliations:** Department of Nursing, Wisconsin International University College, Kumasi, Ghana; Department of Nursing, Garden City University College, Kumasi – Ghana; Faculty of Allied Health, Department of Nursing, Kwame Nkrumah University of Science and Technology, Kumasi, Ghana; Department of Mathematics, Ghana Education Service, Serwaah Nyarko Girls’ Senior High School, Kumasi, Ghana; Surgery Directorate, Komfo Anokye Teaching Hospital, Kumasi, Ghana

**Keywords:** Breast cancer survivors, chemotherapy, radiotherapy, quality of life, mastectomy, Physical Well-being, Pain, Distorted Sleep, Fertility, Fatigue

## Abstract

**Background:** Years back, cancer was thought to be a problem limited to the developed countries. Cancer is now a disease with lots of burden, leading cause of death and disability in developing countries. Physical health is very important for the overall well-being of breast cancer survivors, since it is the visible part of the dimensions of overall health and well-being. Prior studies have examined factors leading to late detection, financial burden and experiences of caregivers of breast cancer patients in Ghana, however, none of the studies have explored how breast cancer may specifically impact the physical well-being and the quality of life of these women. This study therefore seeks to explore how breast cancer impact the physical well-being and the quality of life of these women.

**Methodology:** The study site and setting was the Oncology directorate at Komfo Anokye Teaching Hospital where twelve respondents were recruited for the study. Breast care survivors were purposively sampled and interviewed (face-to-face) to explore how the BC has impacted on their physical wellbeing and their Quality of Life (QoL). Demographic data were obtained before the main interview. The interviews lasted between 30minutes to 45minutes. Data was analysed using thematic analysis of key information by using NVivo data management software.

**Results:** Participants described their physical wellbeing according to the way they encountered the disease from the onset through to the treatment administration. Five sub-themes emerged, namely: Fatigue/bodily weakness: impaired functional ability: pain; distorted sleep; and fertility.

**Conclusion:** Fatigue, impaired functional ability, pain, distorted sleep and fertility were consistent with constructs of the Quality of Life model. These physical symptoms negatively affected the total well-being of the BCSs.

## Introduction

Years back, cancer was thought to be a problem limited to the developed countries. Cancer is now a disease with lots of burden, leading cause of death and disability in developing countries. Among cancers, breast cancer currently is one of the well-known malignancies with high mortality all over the world affecting women in both developed and developing countries [1].

The International Agency for Cancer Research (IARC) in 2018 posited that breast cancer is the fifth leading cause of death with a record of 627,000 representing 6.6% of the world cancer deaths. Over the years, the incidence of breast cancer has increased enormously with 7.5% in 1975, 12.8% in 1996 and a staggering 12.8% in 2009 for all cancer cases [2, 3].

In Ghana, breast cancer is a major public health problem and the lead mortality rate in cancer cases although it is the fourth cause of death in cancer cases worldwide [4]. Ghana like other Sub-Saharan countries experiences late report of breast cancer symptoms, lack of access to radiotherapy facility, high economic burden and discontinuing follow-up treatment [5].

The treatment of breast cancer largely depends on the stage of diagnosis. The American Cancer Society categorized stages of breast cancer (from stage 0, stage I, stage II, stage III and stage IV) according to the extent to which the cancer cells have evaded the breast. Treatment options include one or a combination of surgery, radiation therapy, chemotherapy, and endocrine or hormone therapy. However, undergoing any of the cancer treatment comes with its side effects that affect the physical, social, psychological and spiritual well-beings of the individual [6].

Physical well-being is “the ability to perform physical activities and carry out social roles that are not hindered by physical limitations and experiences of bodily pain, and biological health indicators”. Physical health is very important for the overall well-being of breast cancer survivors, since it is the visible part of the dimensions of overall health and well-being [7].

Generally, women with breast cancer may experience some changes in their body because of the signs and symptoms of the disease and treatment regime which is very devastating. These visible changes are concealed by the survivors in ways such as wearing of under clothes and wigs [8]. Fatigue is significant and one of the long-term problems that is inevitable to cancer patients and survivors [9]. Breast cancer survivors in advance stage of the disease experience fatigue as the most troubling symptom, increased fatigue is associated with decreased patients’ functional status and QoL [10]. A study on cancer related fatigue posited that, breast cancer related fatigue goes a long way to interfere with the therapy and hence prolong the duration of the treatment [11].

Sleep quality of breast cancer survivors may be distorted as they go through different treatment procedures. A number of studies on QoL of cancer survivors indicated that distortions of sleep of cancer survivors is a key symptom of the disease and its treatment [12]. Among all cancer survivors, breast cancer survivors have the highest prevalence sleep disturbance and subjective complaints which indicate that breast cancer survivors have poorer sleep quality compared to the other cancer survivors. Sleep disturbances among breast cancer survivors may be attributable to varied interacting factors such as pain, fatigue, adverse effects of treatment such as nausea, vomiting, and psychological causes [13].

Fertility plays a significant role in the quality of life of breast cancer survivors, mainly for those of reproductive age [14, 15]. Breast cancer survival rate continue to increase as a result of the emergence and enhanced technological and medical treatment for breast cancer and an increased public education on breast cancer. As the survival rate rises, so does the importance of starting or expanding the family during and after cancer [16]. Due to the side effects of breast cancer treatment and the trauma associated with the disease, getting pregnant sometimes becomes difficult naturally.

Breast cancer survivors can experience pain when the developing tumour press on the nerves or bone or other organs around the breast. Pain from breast cancer can be mild to severe depending on the survivor and extent of the cancer. According to the ACS, peripheral neuropathy, mouth sores and radiation mucositis are common pain experienced by breast cancer survivors during and after chemotherapy and radiation therapy respectively [6]. Breast cancer survivors who had undergone mastectomy might experience chronic, progressively debilitating neuropathic pain in the arm axilla, chest wall and breast [17]. This pain makes it difficult for the patients to perform their routine daily activities which negatively affects their quality of life.

Prior studies have examined factors leading to late detection, financial burden and experiences of caregivers of breast cancer patients in Ghana, however, none of the studies have explored how breast cancer may specifically impact the physical well-being and the quality of life of these women. This study therefore seeks to explore how breast cancer impact the physical well-being and the quality of life of these women.

## Methodology

The study site and setting was the Oncology directorate at Komfo Anokye Teaching Hospital (KATH). The study population included every breast cancer survivor for 6 months and more after being diagnosed of breast cancer, who is eighteen (18) years or more than eighteen years up to 65 years, and who had sought for clinical or non-clinical service(s) at the oncology directorate at KATH as at the time of the study. However, breast cancer survivors who are too sick to communicate and those who are not mentally sound were excluded from the study. Certificate of Registration (RD/CR20/049) was obtained from the Research Unit of the Komfo Anokye Teaching Hospital. In addition, ethical clearances were obtained from Kwame Nkrumah University of Science and Technology Committee on Human Rights Publication and Ethics (CHRPE) with ref no: CHRPE/AP/177/20. The research data was collected from 1st October, 2020 to 31st January, 2021, which falls within the approval period stated in the certificate of registration by the Oncology Unit. This is because no new information was gathered during the interview sessions after the twelveth (12^th^) participant.

After the researcher had obtained the required ethical approvals for this study, BCSs were approached for data collection. The researcher with the help of a senior nursing officer and a staff nurse at the Oncology directorate recruited the respondents who were willing to share their experiences about the BC and its impartation on their physical wellbeing after considering the study’s inclusion and exclusion criteria [18]. The researcher explained the reason for conducting the study to the identified BCSs. An appointment was booked with each participant for an interview at their convenience. A consent form was given to each participant to read and sign before they were interviewed. Respondents for the study were not forced to participate but voluntarily accepted to take part in the study.

The qualitative data collection technique used in collecting data for this study were a semi-structured and unstructured interview. In addition, face-to-face approach was used to administer the data instruments which helped to obtain accurate screening of respondents. The participants recruited for this study were given a consent form to read and sign. The Principal Investigator (PI) informed the participants of their reserved right to withdraw from the study at any point in time they wished to do so. Demographic data were also obtained before the main interview.

The interviews lasted between 30minutes to 45minutes. The interviews were audio recorded with an electronic audio recorder after participants consented to it. Field notes were taken in order not to forget the poster, tone, gestures and signs of each participant. In analysing the data for this study, the researcher transcribed audio-recorded interviews verbatim as a word-processed text. Data was transcribed with each interview recording saved as a separate word-processed file with a codified pseudonym in order to maintain confidentiality and preserve anonymity of the participants. The transcribed data was analysed using thematic analysis of key information by using NVivo data management software.

## Results

In all, 12 women living with breast cancer were recruited for this study. Their ages range from 30 years to 62 years. All participants had some form of education, where two of them had primary education, six of the participants completed their secondary education and the remaining four had tertiary education. Seven of the participants were married, one was divorced before diagnosis, one was widowed, two were unmarried and one has been separated from the husband after her diagnosis. One of the participants became unemployed after the diagnosis, one was a student, and two of them were on retirement. The diagnosis of the disease and side effects of its treatment forced one of the two pensioners to go on voluntary retirement. The remaining eight of the participants were gainfully employed. Their occupation included trading, banking, accounts clerk, nursing and teaching. All the participants were Christians with the exception of two participants who were Muslims.

In addition, all the twelve participants had undergone some form of cancer treatment. Eleven of the participants were receiving chemotherapy and a participant was on injection Faslodex.

Among the participants who were receiving chemotherapy, three had undergone surgery (mastectomy). Only one of the participants had received chemo therapy, surgery and radio therapy. The participants have been living with the disease for 8 months to 5 years. Again, all the twelve participants had children: two had a child; three had 2 children; two had 3 children; four had 4 children; and one had 5 children.

### Physical Well-being

Participants described their physical wellbeing according to the way they encountered the disease from the onset through to the treatment administration. Five sub-themes emerged, namely: Fatigue/bodily weakness: impaired functional ability: pain; distorted sleep; and fertility.

### Fatigue/Bodily Weakness

Fatigue is major symptom of breast cancer and one of the components of the physical well-being of the QoL model. Most of the participant disclosed that they experienced fatigue/bodily weakness at a point in the trajectory of the disease. Two of the participants narrated that, they felt tired and general body weakness that led them to report to the hospital for the detection of breast cancer.

> *“I was in my usual state of health until about 7 months ago when I started feeling generalized body weakness, I came to the poly clinic, narrated what brought me there and was referred to the BCC after which some laboratory investigations and mammogram were done and I was later referred to the oncology directorate.”* (**Cynfo)**

> *“I used to walk about 2 kilometres to the road side every morning to board ‘trotro’ or taxi to the workplace. But now, I have to get 4 or 6 rests in between the same distance before I reach the roadside. I easily get tired when I do little work.” **(Marzu)***

Some of the women complained of generalized body weakness after receiving the chemo therapy. They usually spend about 7 days to 14 days to recover from the body weakness.

> *“My major concern is the general body weakness after receiving the chemo therapy. Usually, I spent 2 weeks in recovery.” **(Geortim)***

> *“… after I started with the therapy, I became very weak… It takes some weeks to recover. The interval between the cycles is 3 weeks and by the time you recover then you are due for the next cycle.” **(Beakwa)***

### Impaired Functional Ability

The women claimed that, their physical activities have been affected negatively. They could not go to work due to fatigue, body weakness, pain and other side effects from cancer treatment. Even after recovery from the above-mentioned side effects of cancer treatment, they are unable to perform duties like they used to do before they were diagnosed. They lamented;

> *“Yes, it has really affected my work because when I started the chemo, I have not been able to go to work. After the chemotherapy, I cannot do anything on my own especially in the first week after receiving the chemo. I cannot cook, sweep or walk for a distance or even take care of my children.” **(Flochi)***

> *“Initially before I started the therapy, I use to go to work but now I have not been able to go to work. So far, I have received six cycles of chemotherapy but it was after the fourth chemotherapy in which a surgery was performed hence, I have not been able to go to work.” **(Monbe)***

> *“Yes, I felt very weak so I decided not to go to work after the fourth chemo besides I have had a mastectomy done about a month ago hence I am here again for the next cycle of treatment.” **(Maryeng)***

The functional ability of BCS was affected because their work rate had been reduced. One participant reported that she had reduced her workload to the minimal and exercise a little after she was diagnosed of the disease.

> *“I can only perform little daily activities; I spend most of my time in the hospital and at home but I walk every day from my house to the roadside and back to the house.” **(Magdas)***

A participant reported that, she goes to work and do other daily activities after she had recovered from the side effects of the cancer treatment.

> *“I am a trader and do my own work, so I go to work when I have recovered from the therapy.” **(Cynfo)***

Whilst other participants resume work as soon as they have recovered from the side effect of the treatment, one of the women lamented that she cannot go to work again.

> *“Now, I can no longer perform my activities of daily living, I cannot work anymore.” **(Doropo)***

### Pain

Pain was commonly reported by majority of the study participants. Some of the women lamented that their pain resulted from the lump detected in the breast before diagnosis.

> *“I felt some severe pains below my armpit without any associate weight gain/loss, discharges, ulcer or lump three days. I could not raise my hands up and that made me report to the hospital” **(Magdas)***

> *“I was in my usual state of health performing my normal daily routine activities until I started feeling pains in my right breast as a result of an acne under my right breast. I squeezed it out. Few days later I noticed a lump under that same breast without any discharge. It was painful than the previous one, so I reported to the hospital.” **(Dorata)** “At the BCC I felt severe breast pains, a breast biopsy was done before I was referred to the oncology directorate” **(Cynfo)***

For some of the study participants, side effects of chemo and radiotherapy were the major causes of pain experienced. Two of the respondents lamented that;

> *“After every chemotherapy, all my joints start paining me which slows me down.” **(Geortim)***

> *“Aside breast pains, I always had head ache after I had received a chemotherapy. I also experienced pains in my joints but how severe it is do not match up to that of my breast pain and head.” **(Doropo)***

For those who had surgery, the wounds were painful. A participant shared her experience.

> *“The wound after my mastectomy was very painful. My chest area was swollen and I could not touch it. It was not easy…” **(Marzu)***

After series of visit to the oncology directorate at KATH and therapies received, majority participants claimed that their pains had reduced drastically from severity to mild or no pain. Others said that their pains had increased from mild to severe. One of the participants said;

> *“My breast was very painful, tensed and the size had increased without any associated discharges or ulcer. I have been coming for a regular visit to this clinic for almost 3 years now, by the grace of God, I have no pain but sometimes I experienced the tenderness. Yes, it is on/off.” **(Chrisu).***

### Distorted Sleep

The participants reported they were unable to sleep continuously without any interruption emanating from the associated symptoms and treatment side effects. Almost all of the study participants said pain was the main cause of their inability to sleep well. Two of the participants narrated how pain and chemo therapy led them to sleepless times;

> *“I could not sleep for about a month. My breast was painful. When I turn on my bed to a new position, I feel sharp pain in my breast which just wakes me up.” **(Cynfo)***

> *“As I said earlier, it is not easy being on the chemotherapy, you are always weak, you are unable sleep but always drowsy.” **(Beakwa)***

One woman pointed out that, she could not sleep for about 3 months after the doctor diagnosed her of breast cancer. In her sleep, she often got frightened of the disease and woke up. Falling asleep again was difficult for this woman because she had to wait for some minutes.

> *“On the day of hearing my diagnosis, I could not sleep the whole night. I will say I did a wake keeping, hmmm… because I felt asleep around 5:30 am. Afterwards, I just woke up at night frightened and panic over my diagnosis. Going back to bed was difficult for me. It continued for about three months until my third chemo therapy. …I reported to the doctor and I was given some tablets to make me sleep.” **(Chrisu)***

### Fertility

Breast cancer and its treatment can affect the fertility of the survivor. Seven of the participants were in their reproductive age. All participants who were put on chemo therapy were informed by the doctor that their chances of conception would lessen. So, they were advised to wait and complete the chemo therapies before they plan to conceive. One of the participants said;

> *“Before I started the chemotherapy, I was told, “my chances of giving birth again will be 50%” thus, you may complete the chemotherapy and may or may not be able to give birth or conceive again. … I am hoping to give birth again because I have only one child, but I have been advised not to get pregnant whilst receiving the chemotherapy. **(Flochi)***

## Discussion

### Demographic Characteristics of Breast Cancer Survivors

The age distribution of the 12 participants in this study ranged from 30 to 62 years. Ten (10) among the women were above 40 years, and two (2) of them were below 40 years. This gives an indication that breast cancer does rarely affect women under 40 years (CDC, 2019).

Three out of the twelve (12) participants had family history of breast cancer and other cancers. This agrees with past studies conducted which posited that 20% - 30% of newly diagnosed breast cancers are associated with history of cancer in first-degree and history of mammary gland diseases [19].

More so, all the study participants had formal education but completed at different levels. Only two had primary education. The rest of them (10) had secondary education with four (4) among them attained tertiary education. This could be attributed to the urban nature of the study setting tagged as the second most populous and largest city in Ghana. It could also be attributed to the assumption that educated people are more likely to avail themselves for breast screening. This is because lack of education can reduce access to opportunities and health resources [20, 21, 22].

Lastly, all the participants were gainfully employed before their diagnosis. This could be the reason why their BC were detected early since they were able to afford the initial health cost. This finding reiterates a study by Khater and friends who reported that Egyptian women with BC were diagnosed early because they could afford the initial breast cancer detection screening [23].

### Physical Well-being of Breast Cancer Survivors

In this study, the physical well-being of breast cancer survivors was largely associated with the signs and symptoms of the disease and side effects of its treatment. Some of the study participants had moderate or poor experiences on their physical well-being components. In general, the findings from the current study revealed that the participants had poor physical well-being. This reiterates the findings of other studies related to poor physical well-being of BCSs [24, 25]. Fatigue, pain, impaired functional ability and distorted sleep were the common experiences encountered among women with BC as per this study’s findings.

Again, fatigue is a common symptom experienced by women living with BC [9]. This finding is consistent with the findings by Hofman and colleagues who reported that, 70% to 90% of breast cancer survivors reported about experiencing fatigue. This Breast cancer related fatigue (BCRF) may go a long way to interfere with the therapy and hence prolong the duration of the treatment [11]. The study findings revealed that, the participants were willing to carry on with their daily household chores and workplace duties but they could not perform any activity or perform without assistance because they easily get tired. This finding affirms a study by Bovbjerg and friends who posited that fatigue made BCSs unable or less able to perform their daily routine activities. In effect, the quality of life of breast cancer survivors were profoundly and pervasively affected by fatigue as already reported [28].

Pain associated with breast cancer has been investigated by various scholars around the world and reported accordingly [2]. This study findings showed that all the participants experienced pain as one of the main constituents of physical well-being. According to the study participants, the pain level ranged from mild to severe which was the same as the findings in other studies on BC pain [27, 30]. Majority of the study participants claimed that they reported to the hospital after experiencing pain in their breast which led to their diagnosis. This affirms the statement that BC itself often causes pain [31]. Four of the participants who had undergone mastectomy complained of the pain after the surgery and its effect on their livelihood. This finding was also consistent with a study conducted by Jadhav and Poovishnu Devi on post mastectomy syndrome [17]. Although the pain levels of the women were managed to a moderate level, it was difficult for the women to perform their daily activities without assistance. This finding was congruent with other studies which concluded that pain makes it difficult for BCSs to perform their daily activities and increased the impact on QoL [32].

Distorted sleep among the BCSs were found in this study. This is not surprising because a lot of studies had established distorted sleep among BCSs and cancer patients in general [33]. The sleep quality of the study participants was distorted as they went through different treatment procedures and pain. In other words, the women attributed the cause of their distorted sleep to pain and treatment of BC. This finding was in line with that of other studies [12,13]. Sleep disorders experienced by BCSs impacted negatively on their work productivity and medication abuse and misuse, thus affecting their quality of life [34]. In treating the distorted sleep, non-pharmacotherapy such as yoga and acupuncture were not popular, so the participants were put on pharmacotherapy like sleeping pills to enable them sleep well [33].

Breast cancer and its treatment can affect the fertility of the survivor. All participants who were receiving chemo therapy were informed by their doctor that their chances of conceiving would lessen and even if it becomes difficult to conceive, they will be assigned to a specialist to intervene. Seven of the participants who were in their reproductive age were not perturbed because they saw other BCSs who had given birth without breasts. The women were thought that they could give birth again after completing their treatment. The fertility of the study participants did not affect their QoL as reported by other studies that BC and its treatment can affect the fertility of the survivor. [14, 15].

The potential capacity of a BCS to perform her routine activities and tasks could be altered by BC and its treatment. This study’s findings revealed that, all the participants were unable to perform their duties immediately after they have undergone their treatment. It was obvious from the findings that, the participants had to relax for seven (7) or more days to recover before they were able to perform their duties. Some of the BCS had to be assisted by friends and family members to perform their duties even when they had recovered. The participants attributed their inability or difficulty to perform their routine activities to side effects of the chemo therapy such as fatigue, pain, depression and body disfigurement which was not different from the results of existing studies [35, 36]. The functional ability of the study participants was poor especially after they have received a chemo therapy, hence impacting negatively on their QoL.

## Conclusion

Fatigue, impaired functional ability, pain, distorted sleep and fertility were consistent with constructs of the Quality of life model. These physical symptoms negatively affected the total well-being of the BCSs.

## Limitations of the study

The study selected only few participants to provide in-depth information in exploring the physical wellbeing of breast cancer survivors due to the qualitative approach employed which will make the findings difficult to generalise. Further research into the psychological wellbeing of BCSs is recommended.

## Data Availability

Data collected this study are available at https://doi.org/10.6084/m9.figshare.24953595

## Acknowledgement

“With Christ in the vessel I smile at the storm”, my first utmost gratitude goes to the almighty God, for the wisdom, protection and guidance during this work. My heartfelt appreciation goes to to my husband who believed in me and stood for me against all odds to further my education, may God continue to bless you. My appreciation goes to the head and staff of the Oncology Directorate of the Komfo Anokye Teaching Hospital for their support and cooperation during the recruitment stage. My deepest appreciation goes to all breast cancer survivors who availed themselves to participate in this work, without their cooperation this work would not have been successful. And to all my wonderful course mates, I am grateful and happy to have met you all, especially Gideon A. Atanuriba.

## References

1. Bigatti, S. M., Wagner, C. D., Lydon-Lam, J. R., Steiner, J. L., & Miller, K. D. (2011). Depression in husbands of breast cancer patients: Relationships to coping and social support. Supportive Care in Cancer, 19(4), 455–466. 10.1007/s00520-010-0835-8

2. Calys-Tagoe, B. N. L., Senaedza, N. A. H., Arthur, C. A., & Clegg-Lamptey, J. N. (2017). Anxiety and depression among breast cancer patients in a tertiary hospital in Ghana. Education (Highest completed), 19, 12–5.

3. GLOBOCAN (IARC). (2018). Section of cancer information. Retrieved from https://gco.iarc.fr/today/fact-sheets-populations

4. Naku Ghartey Jnr, F., Anyanful, A., Eliason, S., Mohammed Adamu, S., & Debrah, S. (2016). Pattern of Breast Cancer Distribution in Ghana: A Survey to Enhance Early Detection, Diagnosis, and Treatment. International Journal of Breast Cancer, 2016. 10.1155/2016/3645308

5. Bemah, A. B., & Ncama, B. P. (2019). Recognizing and appraising symptoms of breast cancer as a reason for delayed presentation in Ghanaian women: A qualitative study. 1– 22.

6. American Cancer Society, (2016). Understanding a Breast Cancer Diagnosis, Types of Breast Cancer. American Cancer Society. Understanding a Breast Cancer Diagnosis, 1–42. Retrieved from https://www.cancer.org/cancer/breast-cancer/understanding-a-breast-cancer-diagnosis/types-of-breast-cancer.html#references

7. Capio, C. M., Sit, C. H., & Abernethy, B. (2014). Physical well-being. In Encyclopedia of quality of life and well-being research (pp. 4805–4807). Springer.

8. Rasmussen, D. M., Hansen, H. P., & Elverdam, B. (2010). How cancer survivors experience their changed body encountering others. European Journal of Oncology Nursing, 14(2), 154–159.

9. Pleun, J., de Klerk, C., Timman, R., Hinz, A., & van der Rijt, C. C. (2012). Differences in fatigue experiences among patients with advanced cancer, cancer survivors, and the general population. Journal of pain and symptom management, 44(6), 823–830.

10. Dodd, M. J., Cho, M. H., Cooper, B. A., & Miaskowski, C. (2010). The effect of symptom clusters on functional status and quality of life in women with breast cancer. European Journal of Oncology Nursing, 14(2), 101–110.

11. Savina, S., & Zaydiner, B. (2019). Cancer-Related Fatigue: Some Clinical Aspects. Asia-Pacific journal of oncology nursing, 6(1), 7–9. 10.4103/apjon.apjon_45_18

12. Chang, W. P., & Chang, Y. P. (2019). Meta-Analysis of Changes in Sleep Quality of Women with Breast Cancer before and after Therapy. Breast Care. S. Karger AG. 10.1159/000502943

13. Matthews, E. E., Tanner, J. M., & Dumont, N. A. (2016). Sleep disturbances in acutely ill patients with cancer. Critical Care Nursing Clinics, 28(2), 253–268.

14. Deshpande, N. A., Braun, I. M., & Meyer, F. L. (2015). Impact of fertility preservation counselling and treatment on psychological outcomes among women with cancer: a systematic review. Cancer, 121(22), 3938–3947.

15. Goncalves, V., & Quinn, G. P. (2016). Review of fertility preservation issues for young women with breast cancer. Human Fertility, 19(3), 152–165.

16. Dinas K.D. (2020) Fertility Counselling and Preservation for Breast Cancer Patients. In: Alipour S., Omranipour R. (eds) Diseases of the Breast during Pregnancy and Lactation. Advances in Experimental Medicine and Biology, vol 1252. Springer, Cham.

17. Jadhav, K. G., & Devi, T. P. (2020). Impact of Post Mastectomy Pain Syndrome on Health-Related Quality of Life in Modified Radical Mastectomy Patients. Indian Journal of Public Health Research & Development, 11(5), 172–176.

18. Rahi, S. (2017). Research Design and Methods: A Systematic Review of Research Paradigms, Sampling Issues and Instruments Development. Retrieved October 22, 2019, from International Journal of Economics & Management Sciences website: https://www.researchgate.net/profile/Samar_Rahi/publication/316701205_Research_Design_and_Methods_A_Systematic_Review_of_Research_Paradigms_Sampling_Issues_and_Instruments_Development/links/590dde424585159781859d9a/Research-Design-and-Methods-A-Systematic.

19. Kamińska, M., Ciszewski, T., Łopacka-Szatan, K., Miotła, P., & Starosławska, E. (2015). Breast cancer risk factors. Przeglad Menopauzalny. Termedia Publishing House Ltd. 10.5114/pm.2015.54346

20. Street Jr, R. L., Makoul, G., Arora, N. K., & Epstein, R. M. (2009). How does communication heal? Pathways linking clinician–patient communication to health outcomes. Patient education and counselling, 74(3), 295–301.

21. Halbach, S. M., Ernstmann, N., Kowalski, C., Pfaff, H., Pfoertner, T. K., Wesselmann, S., & Enders, A. (2016). Unmet information needs and limited health literacy in newly diagnosed breast cancer patients over the course of cancer treatment. Patient education and counselling, 99(9), 1511–1518.

22. Kugbey, N., Meyer-Weitz, A., & Asante, K. O. (2019). Access to health information, health literacy and health-related quality of life among women living with breast cancer: Depression and anxiety as mediators. Patient education and counselling, 102(7), 1357–1363.

23. Khater, A. I., Noaman, M. K., Abdel Hafiz, M. N., Moneer, M. M., & Elattar, I. A. (2019). Health-Related Quality of Life among Egyptian Female Breast Cancer Patients at the National Cancer Institute, Cairo University. Asian Pacific journal of cancer prevention: APJCP, 20(10), 3113–3119. 10.31557/APJCP.2019.20.10.3113

24. Okoli, C., Anyanwu, S. N. C., Ochomma, A. O., Emegoakor, C. D., Chianakwana, G. U., Nzeako, H., & Ihekwoaba, E. (2019). Assessing the Quality of Life of Patients with Breast Cancer Treated in a Tertiary Hospital in a Resource-Poor Country. World Journal of Surgery, 43(1), 44–51. 10.1007/s00268-018-4772-x

25. Purnama Sari, N. P. W. (2019). Physical Wellbeing in Cervical and Breast Cancer Survivors: A Cross-sectional Study in Surabaya, Indonesia. Indonesian Journal of Cancer, 12(3), 80. 10.33371/ijoc.v12i3.614

26. Hofman, M., Ryan, J. L., Figueroa-Moseley, C. D., Jean-Pierre, P., & Morrow, G. R. (2007). Cancer-related fatigue: the scale of the problem. Oncologist, 12(1), 4.

27. Bovbjerg, D. H., Keefe, F. J., Soo, M. S., Manculich, J., Van Denburg, A., Zuley, M. L., & Shelby, R. A. (2019). Persistent breast pain in post-surgery breast cancer survivors and women with no history of breast surgery or cancer: associations with pain catastrophizing, perceived breast cancer risk, breast cancer worry, and emotional distress. Acta Oncologica, 58(5), 763–768.

28. Hiensch, A. E., Bolam, K. A., Mijwel, S., May, A. M., & Wengström, Y. (2020). Sense of coherence and its relationship to participation, cancer-related fatigue, symptom burden, and quality of life in women with breast cancer participating in the OptiTrain exercise trial. Supportive Care in Cancer, 28(11), 5371–5379. 10.1007/s00520-020-05378-0

29. American Cancer Society, (2019). Treatment and Support. Retrieved from https://www.cancer.org/treatment/survivorship-during-and-after-treatment.html

30. Chapman, E. J., Edwards, Z., Boland, J. W., Maddocks, M., Fettes, L., Malia, C., Bennett, M. I. (2020, April 1). Practice review: Evidence-based and effective management of pain in patients with advanced cancer. Palliative Medicine. SAGE Publications Ltd. 10.1177/0269216319896955

31. Rasmussen, G. H. F., Kristiansen, M., Arroyo-Morales, M., Voigt, M., & Madeleine, P. (2020). Absolute and relative reliability of pain sensitivity and functional outcomes of the affected shoulder among women with pain after breast cancer treatment. PLoS ONE, 15(6). 10.1371/journal.pone.0234118

32. Heary, K. O., Wong, A. W. K., Lau, S. C. L., Dengler, J., Thompson, M. R., Crock, L. W., Novak, C. B., Philip, B. A., & Mackinnon, S. E. (2020). Quality of Life and Psychosocial Factors as Predictors of Pain Relief Following Nerve Surgery. HAND. 10.1177/1558944720911213

33. Otte, J. L., Carpenter, J. S., Manchanda, S., Rand, K. L., Skaar, T. C., Weaver, M., Landis, C. (2015, February 1). Systematic review of sleep disorders in cancer patients: Can the prevalence of sleep disorders be ascertained? Cancer Medicine. Blackwell Publishing Ltd. 10.1002/cam4.356

34. Siefert, M. L., F. Hong, B. Valcarce, & D. L. Berry, (2013). Patient and clinician communication of self-reported insomnia during ambulatory cancer care clinic visits. Cancer Nurs. 37:E51–E59.

35. Modena, C. M., Martins, A. M., Ribeiro, R. B. N., & Almeida, S. S. L. d. (2014). Men and the process of illness by cancer: a look at the Brazilian scientific production. Revista Baiana de Saúde Pública, 37(3), 644–660. Retrieved from http://files.bvs.br/upload/S/0100-0233/2013/v37n3/a4466.pdf.

36. The American Society of Clinical Oncology (2020). Breast Cancers. Retrieved from: https://beta.asco.org.

